# Understanding Health Service Utilisation Patterns for Care Home Residents During the COVID-19 Pandemic using Routinely Collected Healthcare Data

**DOI:** 10.1101/2023.07.11.23292499

**Authors:** Alex Garner, Nancy Preston, Camila Caiado, Emma Stubington, Barbara Hanratty, James Limb, Suzanne Mason, Jo Knight

**Affiliations:** Lancaster Medical School – Lancaster University; School of Health and Related Research – The University of Sheffield; Division of Health Research – Lancaster University; Department of Mathematical Sciences – Durham University; Population Health Sciences Institute – Newcastle University; County Durham and Darlington NHS Foundation Trust

## Abstract

**Background:** Healthcare in care homes during the COVID-19 pandemic required a balance, providing treatment while minimising exposure risk. Policy for how residents should receive care changed rapidly throughout the pandemic. A lack of accessible data on care home residents over this time meant policy decisions were difficult to make and verify. This study investigates common patterns of healthcare utilisation for care home residents in relation to COVID-19 testing events, and associations between utilisation patterns and resident characteristics.

**Methods:** Linked datasets including secondary care, community care and a care home telehealth app are used to define daily healthcare utilisation sequences for care home residents. We derive four 10-day sets of sequences related to Pillar 1 COVID-19 testing; before [1] and after [2] a resident’s first positive test and before [3] and after [4] a resident’s first test. These sequences are clustered, grouping residents with similar healthcare patterns in each set. Association of individual characteristics (e.g. health conditions such as diabetes and dementia) with healthcare patterns are investigated.

**Results:** We demonstrate how routinely collected health data can be used to produce longitudinal descriptions of patient care. Clustered sequences [1,2,3,4] are produced for 3,471 care home residents tested between 01/03/2020–01/09/2021. Clusters characterised by higher levels of utilisation were significantly associated with higher prevalence of diabetes. Dementia is associated with higher levels of care after a testing event, and appears to be correlated with a hospital discharge after a first test. Residents discharged from inpatient care within 10 days of their first test had the same mortality rate as those who stayed in hospital.

**Conclusion:** We provide longitudinal, resident-level data on care home resident healthcare during the COVID-19 pandemic. We find that vulnerable residents were associated with higher levels of healthcare usage despite the additional risks. Implications of findings are limited by the challenges of routinely collected data. However, this study demonstrates the potential for further research into healthcare pathways using linked, routinely collected datasets.

## Introduction

The COVID-19 pandemic had a major impact on adult social care. There was substantial excess mortality in care homes during the first phase of the COVID-19 pandemic, deaths were estimated 20% higher than previous years, a large portion of which are not registered as due to COVID-19^1,2^. The highest proportion of deaths involving COVID-19 of care home residents in wave one was in the North East (30% of deaths involved COVID-19) ^2^. Best policy for care homes was uncertain at the beginning of the pandemic. Studies have shown long-term decline in health related quality of life and functional decline in older patients who were hospitalised for COVID-19^3^. Healthcare for vulnerable people required a fine balance, to ensure necessary healthcare was maintained while minimising exposure to COVID-19 which was particularly pertinent in care homes^4^.

During the early stages of the pandemic, policy recommendations for care homes were updated and revised rapidly. Between the initial COVID-19 guidance on 25th February 2020 and £850m social care grant to councils on 16^th^ April 2020, Public Health England and the Department of Health and Social Care provided numerous additional frameworks and guidance doument^5^. These were often vague and difficult to follow^6^. Criticisms have described the UK’s policy response in adult social care as ‘slow, late and inadequate’^7^.

On 17^th^ March 2020 NHS England advised that all non-urgent elective operations should be postponed, and for all medically fit inpatients to be discharged to free-up capacity^8^. Grimm et al found that care home residents’ use of inpatient care decreased in the early stages of the pandemic and suggest these reductions may result in substantial unmet healthcare need ^9^. In a global survey in the early stages of the pandemic, two-thirds of health care professionals for chronic diseases stated moderate or severe effects on their patients due to changes in healthcare services^10^.

Care home residents have high levels of physical dependency, cognitive impairment, multiple morbidity, and polypharmacy^11^. Comorbidities such as diabetes and dementia are prevalent in the population and require ongoing high levels of care from staff and specialists^12,13^. Dementia was the most common pre-existing condition for residents who died of COVID-19 before the end of 2021 and diabetes was a common comorbidity for male residents who died of COVID-19 in the same period ^2^.

There is a lack of patient-level data from care homes themselves and it is difficult to identify care home residents from administrative hospital data^14^. This limits studies using routinely collected hospital data on care home residents and reduces the possible evidence base for policymakers ^15^.

Using a unique dataset of linked, routinely collected care home and hospital data, we built a description of healthcare of care home residents during the pandemic. We investigate how specific circumstances such as positive COVID-19 impacted care patterns between care homes and the healthcare system to find common groups of longitudinal healthcare utilisation leading to COVID-19 cases, or in response to COVID-19 tests. We provide an investigation into patterns of care and possible associations with resident characteristics and outcomes.

## Methods

### Data Source

We utilised data from the *HealthCall Digital Care* Homes app that began rollout in the North East of England 3^rd^ August 2018. HealthCall is a digital referrals app used by care home staff to gather information and request review from a clinician. Three care home datasets from HealthCall covering resident enrolment, home enrolment, and app uploads are linked via pseudonymised NHS number to eight routinely collected datasets from County Durham and Darlington NHS Foundation Trust hospitals (CDDFT), including A&E, inpatient, outpatient, and community data (primary care data is not included). Pillar 1 COVID-19 testing in the region is also included. In total eight of the datasets refer to patient healthcare events. Three datasets include additional information about residents and homes. A description of each dataset is contained in the supplementary materials.

The COVID-19 testing data used for this analysis is Pillar 1 PCR test results. Pillar 1 testing is classed as ‘swab testing in Public Health England (PHE) labs and NHS hospitals for those with a clinical need, and health and care workers’^16^. The testing data consists of tests when a resident is an inpatient, or when a resident is symptomatic or believed to have been exposed to someone with suspected COVID-19.

### Dataset Descriptive Statistics

Monthly numbers of observations are calculated for each of the datasets. Locations of COVID-19 tests and rates of test results at the different location types were calculated and independence of these two factors was tested with a chi-squared test (see supplementary material).

### Defining Cohort and Trajectories

Since the data contains the healthcare interactions of all CDDFT service patients, a cohort of care home residents was defined. Presence of individuals’ NHS numbers in the HealthCall enrolment (activation) dataset indicate care home residency. Observations in other datasets referring to a resident living in the set of HealthCall care homes are used to identify additional care home residents. Residents are included in the study from the identified timepoints at which they became a care home resident to when they died or moved out of the home. All individuals identified as care home residents are included in the cohort. Resident characteristics such as age, gender and comorbidities are also drawn from the available datasets (see supplementary material for methods and limitations).

We define a resident’s healthcare trajectory as the sequence of care they received each day. To ensure only one state per day, we prioritise more ‘significant’ types of care. The possible states (in order of significance) are:

- A&E attendance
- Inpatient stay in hospital
- Outpatient attendance
- Appointment in the community
- Care home visit by community healthcare staff
- Care Home – no actions in the datasets

### Sequence Analysis

Four different 10-day sub-sequences of resident trajectories were investigated using index events defined by the available COVID-19 tests. The two index events used are a resident’s *first COVID-19 test* and a resident’s *first positive COVID-19 test*. The sequence length of 10 days corresponds to the UK government recommended isolation period for individuals who test positive for the majority of the study period. Residents without a COVID-19 test were not included. Sequences exceeding the boundaries of the study period or a resident’s time in the cohort were excluded from the analysis.

Pairwise distances were calculated between sub-sequences in each of the four sets using the Optimal Matching distance algorithm^17^. Insertion and deletion costs of 1 were used, and substitution costs were based on the transition rate between the two states (see supplementary materials for more information). The sequences were clustered based on the calculated dissimilarity between them using hierarchical clustering and Ward’s criterion. State Sequence Analysis was implemented in *R* using the *TraMineR* package^18^.

Potential associations between cluster assignment and resident characteristics were investigated to provide insight into which factors are associated with the care a resident received. Specific characteristics were investigated: 28-day mortality after the COVID-19 test and Charlson Comorbidity Index, as well as the prevalent comorbidities: diabetes and dementia. Additional associations with wave of the pandemic and COVID-19 test result are included in the supplementary materials.

Chi-squared tests for independence were used for each of the characteristics separately (or Fisher’s exact test when counts in the elements of the table are ≤ 5)^19^, with an adjusted significance level *α*=0·00143 as a simple Bonferroni multiple testing correction from α = 0·05 (total number of tests presented in the main paper and supplementary materials = 45, 16 are included in the main paper).

### Cluster Transitions

Since the sequences defined lead to, and follow on from, index events we use Sankey diagrams to visualise the movement between clusters.

## Results

8,702 care home residents were identified from 122 care homes. *Table 1* provides a summary of the cohort demographic information.

**Table 1:** Formatted summary table. Includes characteristics of 8,702 identified care home residents * We do not have age information for 1,394 of the residents. ** We could not calculate a Charlson Comorbidity Index for 4,671 residents due to them not having registered ICD-10 codes from their inpatient stay. Percentages are of those calculated.

|  | Median |  | IQR |  |
| --- | --- | --- | --- | --- |
| Age * | 85 |  | 79-90 |  |
| Number of Observations | 58 |  | 29-109 |  |
| Months in the cohort | 19 |  | 11-31 |  |
|  | Male |  | Female |  |
| Gender | 3,086 (35%) |  | 5,616 (65%) |  |
|  | True |  | False |  |
| Died (within the study period) | 2,549 (29%) |  | 6,153 (71%) |  |
|  | 0 | 1-2 | 3-4 | ≥5 |
| Charlson Comorbidity Index ** | 324 (8%) | 2,111 (52%) | 1,292 (32%) | 3-4 (8%) |

*Table 2* summarises 11 datasets, consisting of routinely collected data. The data comes from the CDDFT’s secondary care, community database, observations taken inside the care home on the HealthCall app, and COVID-19 testing data. This data includes residents in the study cohort.

**Table 2:**
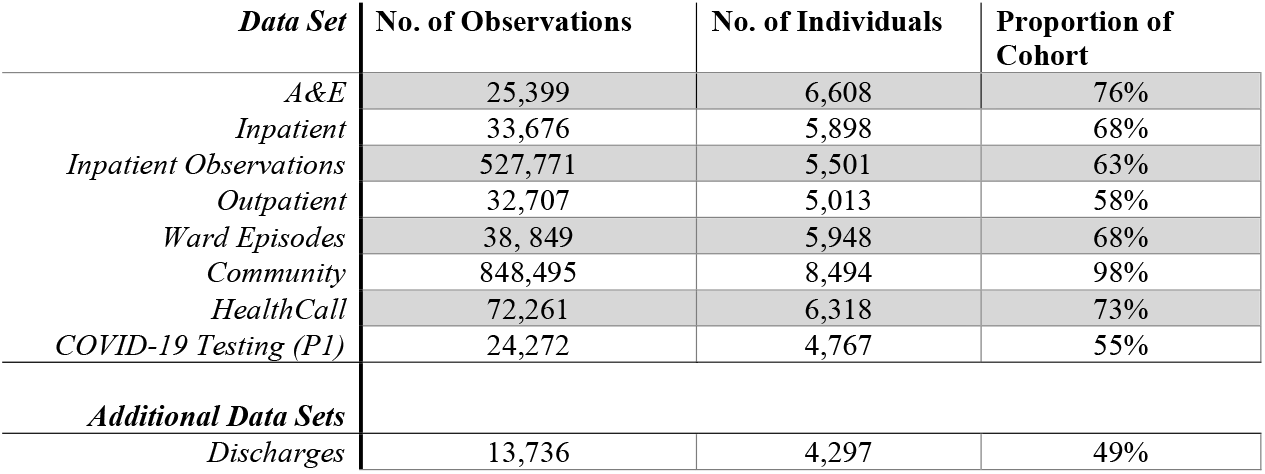
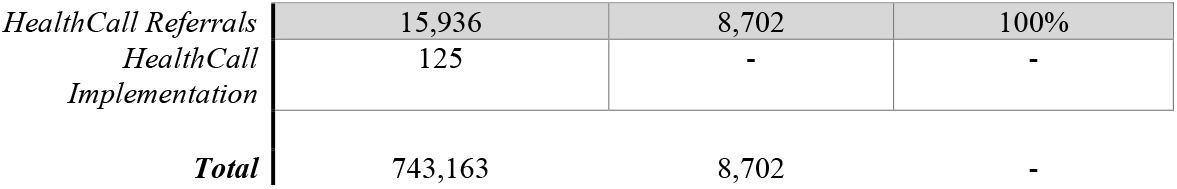
Counts of observations and individuals in each data set, filtered for the cohort of care home residents. * Individuals can be in more than one dataset hence the sums do not equal the total.

Trajectories were defined from the set of healthcare interactions included in the dataset. Figure 1 visualises a resident’s care trajectory throughout their time in the study cohort. The longer blue periods represent an inpatient stay.

**Figure 1:**
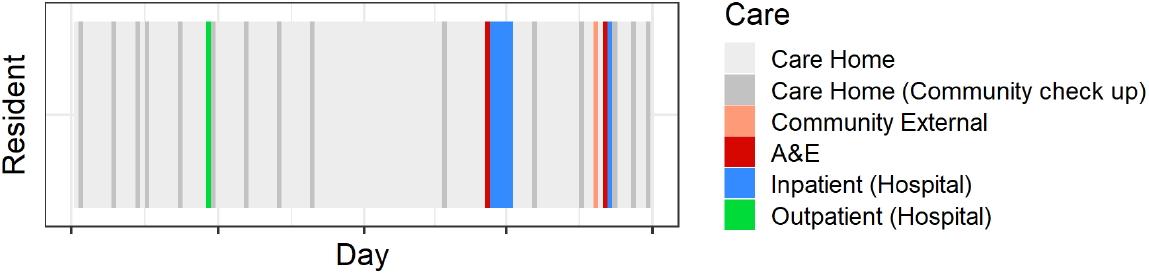
A 5-month sample of a single resident’s care trajectory, with coloured blocks for each day representing the care the resident received each day.

Sequences for clustering were specified based on the COVID-19 testing index events. 4,767 residents have a recorded Pillar 1 COVID-19 PCR test in the dataset, and are therefore included in the analysis, 3,938 were ineligible for analysis due to no testing events. Of these, 1,049 residents test positive for COVID-19 at some point in time and their first tests are used as the index events for the pair of sequences before and after a first *positive* COVID-19 test.

Sequences before the test are not included when a resident moves into the home in the 10 days before the test (198 removed before first positive test, 1,296 removed before a first test). Sequences after the test are not included when the resident dies in the 10-days after the test, or their test is less than 10 days before the end of the study period (316 removed after a first positive test, 1,547 removed after a first test). The number of residents included for each sequence specification is [1] before a first positive test - 851, [2] after a first positive test - 733, [3] before a first test – 3,345, [4] after a first test – 3,220. The total number of individual residents that appear in the analysis is 3,471.

A visualisation of the four 10-day sequences in their assigned clusters can be seen in *Figure 2*. The clusters are generally characterised by a single state. Sequences both before and after the first positive test [1,2] are demonstrated by two clusters: an *inpatient* cluster, and a *home* cluster. The before and after first test sequences [3,4] are characterised by three clusters each, *home, community*, and *inpatient* states and *home, inpatient to home transfer* and *inpatient* sequences respectively. The large number of residents in the *inpatient* cluster after the first test is likely due to testing upon hospital admission. The inclusion of an *inpatient to home transfer* cluster after a first test may indicate that these tests were testing on discharge from the hospital.

**Figure 2:**
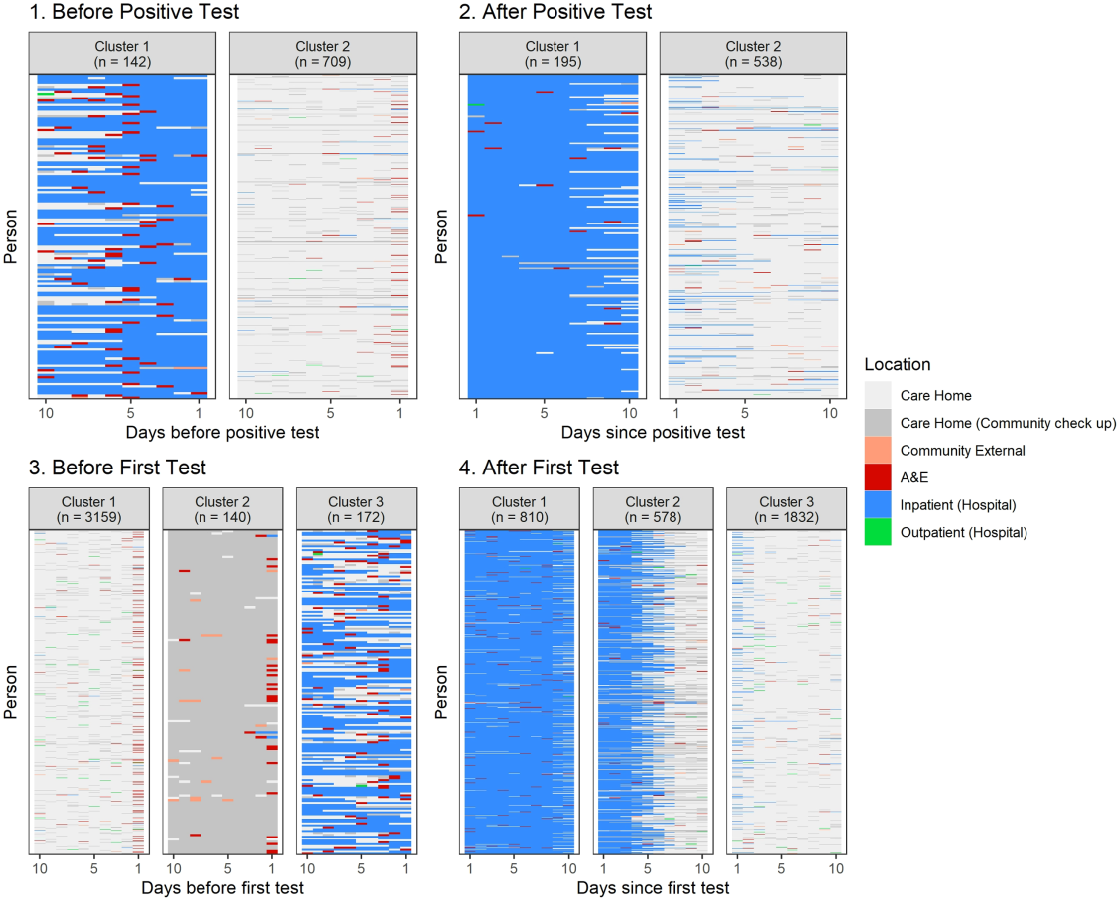
Sequence cluster assignments representing types of care received in the 10 days before (1) and after (2) a resident’s first positive COVID-19 test, and the 10 days before (3) and after (4) a resident’s first COVID-19 test (of any result). The clusters represent the groups of similar sequences, where each sequence represents one resident’s care over the 10 days.

Characteristics of the residents in these clusters were assessed. The relative frequencies of the characteristics within each of the clusters can be found in *Table 3*. The combinations found to be non-independent through the chi-squared test are highlighted in grey. All p-values for these tests can be found in the supplementary materials. A higher proportion of residents with diabetes are found in clusters indicating a higher level of care for all four sequences ([1] p=0.00026, [2,3,4] p<0.0001). For example in the 10 days before a resident’s first positive test 35% are diabetic of 142 in the *inpatient* cluster compared to 21% of 709 in the *home* cluster. A similar pattern is found after both all and positive tests for dementia patients ([2] p=0.00036, [4] p<0.0001). Before all first tests a higher proportion of those in the *community* cluster have frailty scores of 3 and above (73% of 140 versus 41% and 43% for 3,159 in the home and 172 in the inpatient cluster respectively).

**Table 3:**
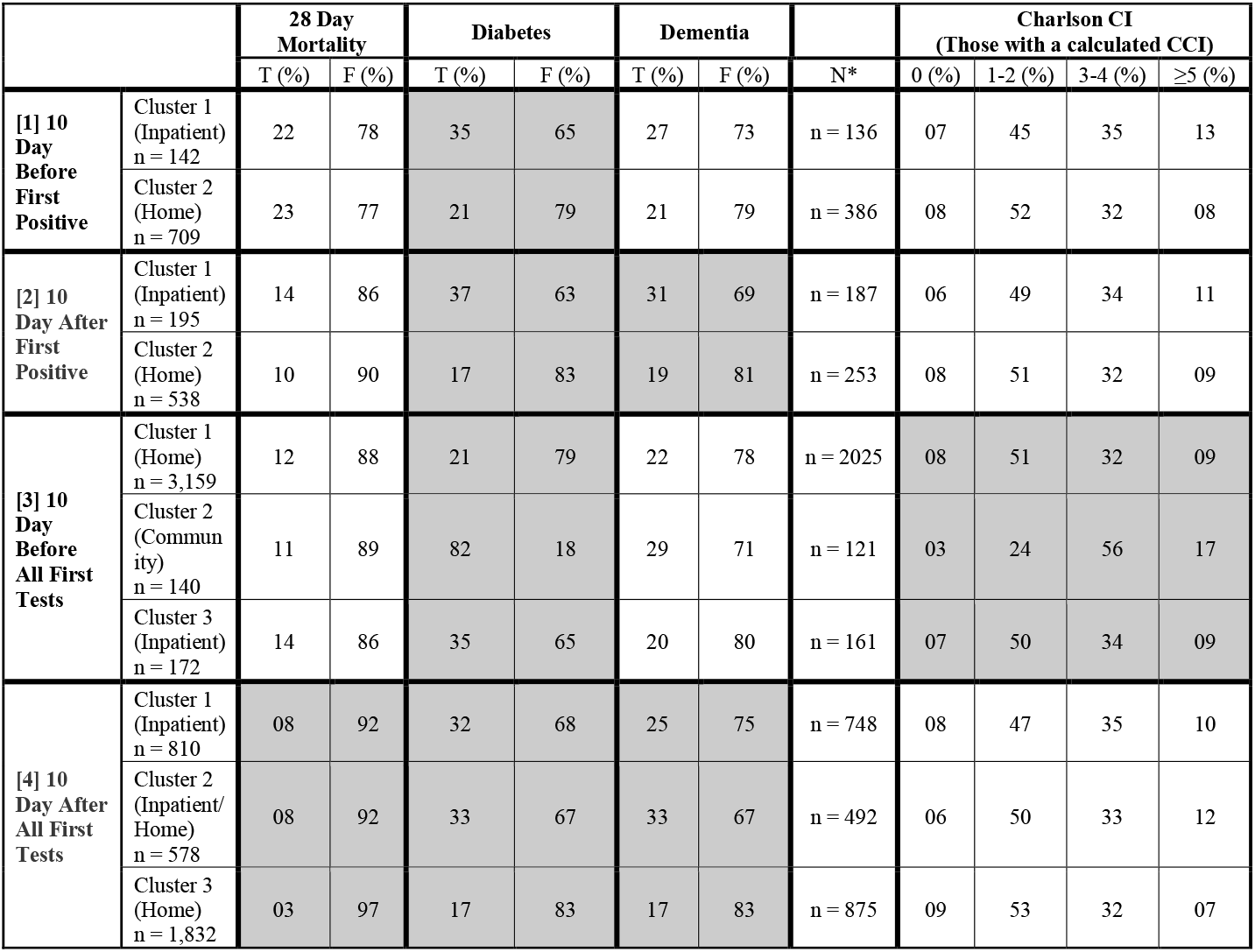
Table of associations between cluster assignments for each of the sub-sequence groups and resident characteristics/sequence outcomes. *Since a Charlson Comorbidity index could not be calculated, we did not include those residents in the proportions and association calculations. The number of residents with a calculated Charlson Comorbidity Coefficient in each group can be seen in the ‘N’ column.

Twenty-eight-day mortality is only associated with clusters 10 days after all tests ([4] p<0.0001); residents in the *inpatient* and *inpatient transfer* cluster have a slightly higher 28-day mortality than those in the *home* cluster (8% of 810 and 8% of 578 versus 3% of 1,832). The two clusters with inpatient stays have the same 28-day mortality rate, despite one of the clusters demonstrating a discharge from hospital around halfway through the 10-day period ([4] p<0.0001).

Flow between clusters before and after the positive test were displayed in a Sankey diagram (*Figure 3*). Transitions between these clusters may indicate changes in care based on the positive test. The ‘*Died*’ after test group here is not the same as presented in the cluster associations previously. Here we identify whether they died within 10 days of their test and were therefore not included in any of the clusters.

**Figure 3:**
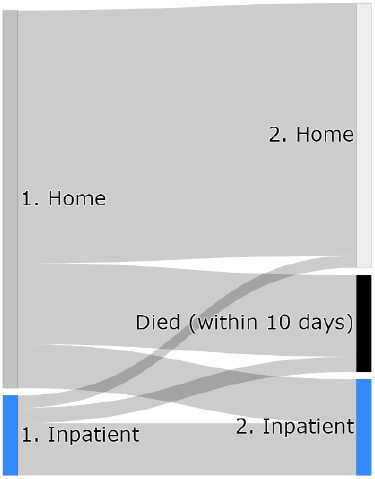
Sankey diagram demonstrating flow between states before and after a resident’s first COVID-19 positive test.

The majority of residents both start and end in the care home cluster. More die within 10 days than are transferred to a stay in hospital. A similar proportion from inpatient care and care homes died within 10 days. This finding could also be an artefact of the usage of Pillar 1 testing data, providing a sample of positive tests that are more likely to be symptomatic in care homes and more routine in hospitals. Alternatively, it may suggest that more residents in the care home should receive hospital care, but also could suggest that the level of care in hospital is not an improvement. We do not account for how ill a resident is, so this could play a part in increasing inpatient mortality rates.

## Discussion

For care home residents the common patterns of healthcare before and after a positive Pillar 1 COVID-19 test generally consisted of residents who stayed in the care home for the whole sequence duration, and those who had the entire duration in hospital. The clusters of healthcare before any first COVID-19 test contain an additional group of residents receiving regular community care across the 10-days before. Clusters after first COVID-19 tests included an additional group of residents who were discharged halfway through the sequence.

Diabetes was always associated with clusters representing higher levels of care. Dementia is associated with inpatient care after a testing event and appears to be highly correlated with a short-term discharge from hospital. Residents who were discharged from inpatient care during the 10-days after their first test appeared to have a similar 28-day mortality rate than those who stayed in hospital.

NHS secondary care use fell during the pandemic. However, the cluster assignments for all of the sequences of care before and after COVID-19 tests and positive COVID-19 tests contain a substantial specific inpatient cluster. There was still a group of residents in hospital, despite the decrease in secondary care use for care home residents at the start of the pandemic^20^.

Dementia is associated with the cluster assignments in the ‘*after*’ event cluster assignments. After the tests there are more residents with dementia in the clusters characterised by the inpatient state, in both the ‘positive tests’ and ‘all tests’ cases, indicating as significant proportion of residents with dementia have transferred into hospital after their test. Residents with dementia are most often in the *inpatient to home transfer* cluster after a first test, which implies that residents with dementia may be more likely to have a shorter stay in hospital. Deciding whether to send residents with dementia for an inpatient stay may be difficult; studies indicate that hospitalisations can be detrimental for individuals with dementia as evidence suggests they are linked with advanced stage of dementia and deterioration of active daily living, among other factors ^21,22^. Evidence suggests that residents with dementia were challenging to care for during the pandemic due to difficulties in adhering to social distancing in both the care home and hospital setting, this may have led to increased hospitalisation as well as high levels of discharge back into homes ^23^.

We provide, to our knowledge, the first in-depth investigation into healthcare patterns of care home residents during the COVID-19 pandemic. Other research provides information on care in the homes during the pandemic, such as that done by Shallcross et al investigating care home-level risk factors among other work^24^. Our findings can be used in context with research on other aspects of residents’ care during the pandemic, to provide thorough policy guidelines for caring for this vulnerable group of residents.

One of the strengths of this study is the unique dataset allowing visualisation and analysis of healthcare for care home residents during the COVID-19 pandemic. Data from community care captures much home-based care, but the absence of primary care data means that some information is absent. We have derived some resident characteristics from secondary and community care history and our record of age and gender is incomplete. Diabetes and dementia are drawn from diagnosis codes for hospital stays and community procedures, hence we are likely to identify the residents who have more advanced disease or who have accessed external care. This is particularly pertinent in the case of dementia, as hospital admission is more likely to be for management of co-occurring conditions ^25^.

A further limitation is that the COVID-19 testing data contains only Pillar 1 tests processed in the Trust’s hospital labs. This may bias the sequences we define (relating to a resident’s first positive COVID-19 test and first COVID-19 test in general), since a large portion of Pillar 1 testing was testing on admission to hospital. Testing outside of hospitals was for those with a clinical need, and are therefore more likely to be tests for symptomatic residents^16^. This is the case when looking at test result rates for the different testing locations, with tests in care homes much more often positive than those in hospital settings (see supplementary materials for breakdown). We find a large portion of the residents in inpatient care before their first positive test, remain in inpatient care afterwards – suggesting COVID-19 may not have been the reason for their admission, but tested positive on arrival. The location of testing differs between wave 1 and wave 2 of the pandemic, we investigated breaking down the clustering analysis into the two waves and found it did not significantly impact the results (both in supplementary materials). The use of Pillar 1 COVID-19 testing allows a consistent level of testing throughout the pandemic, since Pillar 1 testing was introduced first and was conducted over the whole pandemic period. However, a more complete – routine set of COVID-19 tests would give a more accurate description of how residents were treated in general and would allow us to identify residents’ first test and positive test more reliably.

Health services such as the National Health Service of the United Kingdom have large pools of untapped data that can be used for large scale, impactful analyses^26^. Research such as this work is needed to demonstrate the work that can be done going forward using linked, routinely collected datasets. Implications from this study are limited by the nature of Pillar 1 COVID-19 testing. However, this study demonstrates the potential for large scale linkage of routinely collected healthcare data to investigate longitudinal pathways of care for future studies going forward.

## Data Availability

Data was collected from CDDFT and stored in a Trusted Research Environment (TRE) managed by Durham University. Informed consent was not possible as the data was anonymised. The Trust shared anonymised data after undertaking a Data Privacy Impact Assessment and a Data Transfer Agreement. Data supporting this study is not publicly available due to ethical considerations around accessing linked patient level healthcare data. The authors can no longer access the data used in this analysis. Please contact the main author for more information.

## Acknowledgements

Rachel Stocker – PPIE facilitation,

PPIE Group – Helpful Input

Graham King, Catherine McShane, Del Jones, Ian Dove – Assistance of understanding the HealthCall data

## Ethical Approval

The project was approved by Lancaster University Faculty of Health and Medicine Research Ethics committee, reference FHM-2022-3318-RECR-2.

## Declaration

### Ethics approval and consent to participate

The project was approved by Lancaster University Faculty of Health and Medicine Research Ethics committee, reference FHM-2022-3318-RECR-2. Data was collected from CDDFT and stored in a Trusted Research Environment (TRE) managed by Durham University. Informed consent was not possible as the data was anonymised. The Trust shared anonymised data after undertaking a Data Privacy Impact Assessment and a Data Transfer Agreement. There are no specific methodological guidelines for the exploratory work presented in this manuscript.

### Consent for publication

Not applicable.

### Authors’ contributions

J.K. and S.M. acquired the funding.

A.G., J.K, S.M. conceptualised the project.

A.G. cleaned the data.

A.G. performed the analysis with supervision from J.K., N.P., S.M., C.C..

B.H., E.S. and J.L. provided input on analysis and results.

All authors reviewed the manuscript.

## References

1. Hollinghurst J, Lyons J, Fry R, et al. The impact of COVID-19 on adjusted mortality risk in care homes for older adults in Wales, UK: a retrospective population-based cohort study for mortality in 2016–2020. Age and Ageing. 2021;50(1):25–31. doi:10.1093/ageing/afaa207

2. Deaths involving COVID-19 in the care sector, England and Wales - Office for National Statistics. Accessed July 21, 2021. https://www.ons.gov.uk/peoplepopulationandcommunity/birthsdeathsandmarriages/deaths/articles/deathsinvolvingcovid19inthecaresectorenglandandwales/deathsregisteredbetweenweekending20march2020andweekending2april2021

3. Walle-Hansen MM, Ranhoff AH, Mellingsæter M, Wang-Hansen MS, Myrstad M. Health-related quality of life, functional decline, and long-term mortality in older patients following hospitalisation due to COVID-19. BMC Geriatrics. 2021;21(1):199. doi:10.1186/s12877-021-02140-x

4. The Health Foundation. Adult social care and COVID-19 after the first wave: assessing the policy response in England - The Health Foundation. Accessed February 2, 2022. https://www.health.org.uk/publications/reports/adult-social-care-and-covid-19-after-the-first-wave

5. The Health Foundation. COVID-19 Policy Tracker. Accessed June 2, 2022. https://covid19.health.org.uk/home

6. Spilsbury K, Devi R, Griffiths A, et al. SEeking AnsweRs for Care Homes during the COVID-19 pandemic (COVID SEARCH). Age and Ageing. 2021;50(2):335–340. doi:10.1093/ageing/afaa201

7. Daly M. COVID-19 and care homes in England: What happened and why? Social Policy & Administration. 2020;54(7):985–998. doi:10.1111/spol.12645

8. NHS England and NHS Improvement. NEXT STEPS ON NHS RESPONSE TO COVID-19. Published online March 17, 2020.

9. Grimm F, Hodgson K, Brine R, Deeny SR. Hospital admissions from care homes in England during the COVID-19 pandemic: a retrospective, cross-sectional analysis using linked administrative data. Int J Popul Data Sci. 5(4):1663. doi:10.23889/ijpds.v5i4.1663

10. Chudasama YV, Gillies CL, Zaccardi F, et al. Impact of COVID-19 on routine care for chronic diseases: A global survey of views from healthcare professionals. Diabetes Metab Syndr. 2020;14(5):965–967. doi:10.1016/j.dsx.2020.06.042

11. Gordon AL, Franklin M, Bradshaw L, Logan P, Elliott R, Gladman JRF. Health status of UK care home residents: a cohort study. Age and Ageing. 2014;43(1):97–103. doi:10.1093/ageing/aft077

12. Diabetes UK. Good Clinical Practice Guidelines for Care Home Residents with Diabetes.; 2010. Accessed June 14, 2022. https://www.diabetes.org.uk/resources-s3/2017-09/Care-homes-0110_0.pdf

13. Alzheimer’s Society. Home from home: A report highlighting opportunities for improving standards of dementia care in care homes. Published online 2007. https://www.alzheimers.org.uk/sites/default/files/migrate/downloads/home_from_home_full_report.pdf

14. Schultze A, Bates C, Cockburn J, et al. Identifying Care Home Residents in Electronic Health Records - An OpenSAFELY Short Data Report. Published online April 27, 2021. doi:10.12688/wellcomeopenres.16737.1

15. Hanratty B, Burton JK, Goodman C, Gordon AL, Spilsbury K. Covid-19 and lack of linked datasets for care homes. BMJ. 2020;369:m2463. doi:10.1136/bmj.m2463

16. Department of Health and Social Care. Coronavirus (COVID-19) - Scaling up our testing programmes. Published online April 4, 2020. https://assets.publishing.service.gov.uk/government/uploads/system/uploads/attachment_data/file/878121/coronavirus-covid-19-testing-strategy.pdf

17. Abbott A, Forrest J. Optimal Matching Methods for Historical Sequences. The Journal of Interdisciplinary History. 1986;16(3):471–494. doi:10.2307/204500

18. Gabadinho A, Ritschard G, Müller NS, Studer M. Analyzing and Visualizing State Sequences in R with TraMineR. Journal of Statistical Software. 2011;40(1):1–37. doi:10.18637/jss.v040.i04

19. McHugh ML. The Chi-square test of independence. Biochemia Medica. 2013;23(2):143–149. doi:10.11613/BM.2013.018

20. Grimm F, Hodgson K, Brine R, Deeny SR. Hospital Admissions From Care Homes in England During the COVID-19 Pandemic: A Retrospective, Cross-Sectional Analysis Using Linked Administrative Data. Published online February 25, 2021. doi:10.20944/preprints202102.0593.v1

21. Lehmann J, Michalowsky B, Kaczynski A, et al. The Impact of Hospitalization on Readmission, Institutionalization, and Mortality of People with Dementia: A Systematic Review and Meta-Analysis. Journal of Alzheimer’s Disease. 2018;64(3):735–749. doi:10.3233/JAD-171128

22. Wolf D, Rhein C, Geschke K, Fellgiebel A. Preventable hospitalizations among older patients with cognitive impairments and dementia. International Psychogeriatrics. 2019;31(3):383–391. doi:10.1017/S1041610218000960

23. Scerri A, Borg Xuereb C, Scerri C. Nurses’ Experiences of Caring for Long-Term Care Residents With Dementia During the COVID-19 Pandemic. Gerontology and Geriatric Medicine. 2022;8:23337214221077790. doi:10.1177/23337214221077793

24. Shallcross L, Burke D, Abbott O, et al. Factors associated with SARS-CoV-2 infection and outbreaks in long-term care facilities in England: a national cross-sectional survey. The Lancet Healthy Longevity. 2021;2(3):e129–e142. doi:10.1016/S2666-7568(20)30065-9

25. Sommerlad A, Perera G, Singh-Manoux A, Lewis G, Stewart R, Livingston G. Accuracy of general hospital dementia diagnoses in England: Sensitivity, specificity, and predictors of diagnostic accuracy 2008–2016. Alzheimer’s & Dementia. 2018;14(7):933–943. doi:10.1016/j.jalz.2018.02.012

26. Goldacre B, Morely J. Better, broader, safer: using health data for research and analysis.

