## Supplementary Materials Document for "Understanding Health Service Utilisation Patterns for Care Home Residents During the COVID-19 Pandemic using Routinely Collected Healthcare Data"

#### Appendix: Supplementary Materials

#### Methods

| **Healthcare Interaction Datasets** | **Description** |
|---|---|
| A&E | Details of attendances at 5 A&E departments covered by CDDFT including the two major acute hospitals: Darlington Memorial Hospital and University Hospital of North Durham. Date and location of attendance is included, along with details of investigative procedures carried out on the patient and diagnosis codes. |
| Inpatient | Details of inpatient spells in the CDDFT hospitals. Dates for duration of overall stay and ward episodes within the stay are included. ICD-10 (International Statistical Classification of Diseases and Related Health Problems 10$^{th}$ Revision) codes detailing diagnosis and comorbidities. |
| Inpatient Observations | Early Warning Scores of inpatients during their hospital stay (no constituent vital sign observations). Includes ward code of stay, date and time observation was made. |
| Outpatient | Details of outpatient appointments. Includes date and duration of interaction. Includes specialty of staff responsible for the patient. |
| Ward Episodes | Details of patient ward episodes during their hospital stays. Includes the ward code of the episode. |
| Community | Details of community appointments and callouts in the County Durham and Darlington area. Date and location type (conducted at patient's home, in community hospital etc) are included, along with care plan details indicating the reason for the interaction. |
| HealthCall | EWS observations of care home residents logged on the HealthCall app by carers. Contains the separate observations that contribute towards calculating an EWS score and the time the observations were taken. |
| COVID-19 Testing (P1) | Pillar 1 ('swab testing in Public Health England (PHE) labs and NHS hospitals for those with a clinical need, and health and care workers' 1), COVID-19 PCR test results from the Trust's Pathology Lab beginning in March 2020. Includes age at date of test and date of test. |
| **Additional Data Sets** | |
| Discharges | Summary dataset of hospital visits, including number of hospital visits and dates of discharge from hospital. Also includes care home (if applicable) of patient mined from hospital records, and date of death (if applicable) contained in hospital records of the patient. Used as a lookup table for patient death dates. |
| HealthCall Referrals | Dates of activation and deactivation of care home residents on the HealthCall system. Activation dates refer to the date they are first put onto the HealthCall system, may be when HealthCall first goes live in the care home, or when the resident first moves to the care home. Conversely, deactivation dates may refer to the date a resident leaves the care home (moves care home or goes back to own accommodation) or dies. The data identifies the most recent care |

| | |
|---|---|
| | home each resident has been assigned to, providing an indicator of each resident's care home. |
| HealthCall Implementation | Dates each HealthCall care home 'went live' and implemented HealthCall. This is the only non-patient level dataset. |

#### Defining Cohort

##### *Identifying Individuals*

Since the datasets contain information on the majority of CDDFT hospital interactions, not just care home residents, we need to define the cohort of care home residents we are investigating. We can use the common set of pseudonymised NHS numbers to identify this set of individuals through all of the available datasets.

We primarily use the HealthCall activations to define the cohort of care home residents, since this contains all residents registered on the HealthCall system.

This activations data will not include any residents that died before HealthCall went live in the home, since they are not registered on the system posthumously. In order to combat this we identify additional residents of the HealthCall homes using their presence in other datasets:

- Hospital discharge data to an associated HealthCall care home
- COVID-19 tests in HealthCall care homes (over 65s)

We do not include any residents in the cohort who do not have any healthcare interactions (no observations in the healthcare interactions datasets) at all after they are identified.

##### *Date of Addition to Cohort*

We identify the date at which a resident can first be confirmed to be a care home resident, and therefore the date they are added to our cohort, by looking for the first date at which they were observed to be in a care home. This may be before HealthCall was introduced in the care home. The date of addition to the cohort is defined as the earliest of the following types of observations:

- COVID-19 tests in care homes
- Activation on the HealthCall system or any HealthCall uploads
- Inpatient discharge to care homes or to address of a care home in discharge dataset
- Inpatient admission from care homes
- Community contact in residential or nursing homes

##### *Date of Death*

In order to identify when a person has died, we use three sources. The main source of death dates is the discharge dataset, which contains details of deaths known to the trust. We also identify death dates as the discharge date of a resident whose discharge method was "Died", or when there was a registered community appointment for which the care plan sub-category was "Verification of death".

##### *Date of Removal from cohort*

Some residents may move out of the area/group of care homes under investigation. Our death data is likely not comprehensive. We can remove residents from the cohort when they are deactivated from the HealthCall system and not reactivated again. If any of their activations have no deactivation date, they are considered still active. Residents who are removed at this point but not considered to have died unless we have a specific date of death. Their date of removal is whichever is earliest of their registered deactivation from the HealthCall system, or their date of death defined previously.

##### *Inclusion in Sequence Analysis*

Sequences are removed when the sequence definition exceeds the boundary of a resident's time in the cohort. Sequences before the test are not included when a resident moves into the home in the 10-days before the test (198 removed before first positive test, 1,296 removed before a first test). Sequences after the test are not included when the resident dies in the 10-days after the test, or their test is less than 10 days before the end of the study period (316 removed before a first positive test, 1,547 removed after a first test).

#### Identifying Resident Conditions

We have identified residents with certain conditions, so we can compare treatment, trajectories, and outcomes of these people.

We have identified:

- Diabetes
    - Community care plan subcategories; 'diabetes care and management ongoing' and 'blood glucose monitoring'
    - Outpatient appointments with staff type 'DIABETIC'
    - Inpatient ICD-10 codes of; E08, E09, E10, E11, E13
- Dementia
    - Inpatient ICD-10 codes of; F00, F01, F03
- Frailty (Charlson Comorbidity Coefficient) – calculated using the 'comorbidity' R package.
    - Inpatient ICD-10 codes

#### Location & Test Result Correlations

Since the COVID-19 testing data is from Pillar 1 testing, we can investigate rates of testing in each location as well as differences in positivity rates between the locations. Using the trajectories defined in the main paper, we observe an individual's highest level of care each day. We link this to the days the residents in the cohort appear in the COVID-19 testing data. We take one test per person per day and link to their activity on the same day.

We also separate out testing in wave 1 and wave 2 (using the ONS estimations of the start and end of each wave), in order to identify any differences in testing at different stages of the pandemic.

As in the main paper, we use a simple Bonferroni multiple testing adjustment to account for the fact that many tests are conducted in this document. We use an adjusted significance level of $\alpha = 0.000862$, from the original value of $\alpha = 0.05$ and accounting for the 58 tests that are calculated in this document.

#### State Sequence Analysis Background

State sequence analysis is a clustering technique that groups similar sequences of states using a dissimilarity measure. State sequence analysis was used in a health setting by Roux et. al. who use sequences to describe treatment of multiple sclerosis patients, with states describing the level of care consumption within a period of time for each patient [17]. Vogt et. al. used the technique for treatment sequences of heart failure patients aiming to identify and describe common ambulatory care pathways between different providers[18]. Vanasse et. al. used multidimensional state sequence analysis to understand healthcare utilisation for COPD patients, aiming to identify any areas of the healthcare systems where healthcare utilisation can be reduced, and patient outcomes can be improved [19].

#### Number of Clusters Selection – Average Silhouette Width

Once we have performed hierarchical clustering on the sequences, we identify most and least similar sequences. In order to select the optimal number of clusters, we use the average silhouette metric to quantify the relative quality of cluster assignments. The average silhouette width compares the dissimilarities of within-cluster sequences and the between-cluster distances for each sequence. Higher average silhouette widths imply more consistent clusters, so we typically take the number of clusters that maximises average silhouette width.

However, this can be a trade-off since more clusters can make results more difficult to interpret. In each clustering, the trade-off between additional clusters and the additional complexity they bring into the results must be assessed. We used the size of the clusters that are created as an additional constraint on the number of clusters selected, since clusters with fewer than 50 sequences were disallowed, and the number of clusters is reduced by 1 if this criterion is met.

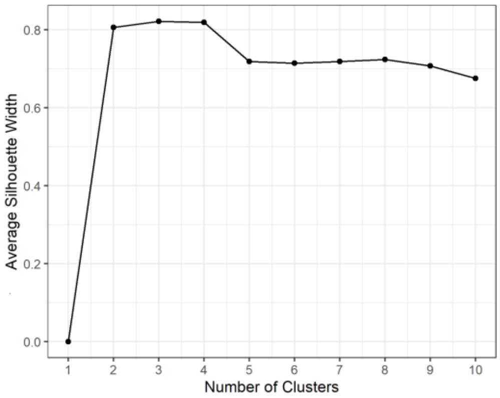


*Figure 1: The average silhouette width for the number of clusters (1-10) in the Before First Positive Test sequences. 2 clusters were selected.*

##### *Optimal Matching*

We use the optimal matching metric to measure similarity (or more accurately, dissimilarity) between two sequences. The optimal matching metric is calculated by transforming one sequence into another, by a sequence of three actions: insertion, deletion, and substitution of states, each with a corresponding cost. The dissimilarity between the two sequences is the (lowest possible) total cost of these actions[2]. More similar pairs of sequences will have smaller a smaller optimal matching metric.

In the case of this study, we set insertion and deletion costs to 1 for any state. The cost of substitution from one state to the other depends on which states are involved in the substitution. We create a symmetrical substitution matrix (n x n), where n is the number of possible states, to define substitution costs. We define the cost of substitution between state A and state B using the transition rate (occurrence of successive states) of state A to state B and state B to state A. States that occur consecutively more often have lower substitution costs. For example, community states are often seen in the middle of care home stays – the community (internal) and care home states often occur consecutively. Therefore replacing a care home state with a community state would likely have a lower substitution cost than the less common A&E state. The maximum possible substitution cost is 2, since this would be the same as deleting state A and inserting state B.

For example, if we have two sequences A & B, both 3 states long. Where A = {CareHome, CareHome, Community} and B = {CareHome, Community, CareHome}. Two ways we could transform B to A are:

1. Insert Care Home as state 1 of sequence B
    a. A = {CareHome, CareHome, Community}
    b. B = {CareHome, CareHome, Community, CareHome}
2. Delete final state of sequence B
    a. A = {CareHome, CareHome, Community}
    b. B = {CareHome, CareHome, Community}

Or

1. Substitute 2$^{nd}$ state of sequence B for CareHome

   a. A = {CareHome, CareHome, Community}
   b. B = {CareHome. CareHome, CareHome}
2. Substitute final state of sequence B for Community
   a. A = {CareHome, CareHome, Community}
   b. B = {CareHome, CareHome, Community}

Since we have more than one way to transform sequence B to sequence A, we choose the lowest cost method. The lowest cost method of transforming sequences is found using the Needleman-Wunsch algorithm[3]. The minimum cost of transforming from sequence A to sequence B is the same as the minimum cost of transforming sequence B to sequence A.

### Results

#### Data Summary

Table 1 shows the datasets used for this study. Numbers of observations are calculated for each of the datasets. Number of individuals is calculated through the number of unique NHS numbers.

*Table 1: Counts of observations and unique individuals in each of the datasets accessed.*

| ***Data Set*** | **No. of Observations** | **No. of Individuals** | **Proportion of Individuals*** |
|---|---|---|---|
| *A&E* | 675,500 | 306,750 | 50% |
| *Inpatient* | 480,745 | 177,403 | 29% |
| *Inpatient Observations* | 3,726,105 | 177,825 | 29% |
| *Outpatient* | 1,770,173 | 328,638 | 54% |
| *Ward Episodes* | 550,358 | 186,885 | 31% |
| *Community* | 3,185,812 | 62,917 | 10% |
| *HealthCall* | 72,261 | 6,318 | 1% |
| *COVID-19 Testing (P1)* | 240,805 | 94,531 | 15% |
| ***Additional Data Sets*** | | | |
| *Discharges* | 47,982 | 20,530 | 3% |
| *HealthCall Referrals* | 15,936 | 8,785 | 1% |
| *HealthCall Implementation* | 125 | - | - |
| ***Total*** | 10,701,759 | 612,408 | - |

Table 1 Legend: * Individuals can be in more than one dataset hence the sums do not equal the total.

#### Cohort Growth

Since our cohort is identified observationally –as time goes on from the start of the study data, there are more data points available to identify our cohort with and the number of residents who fit the criteria increases over time.

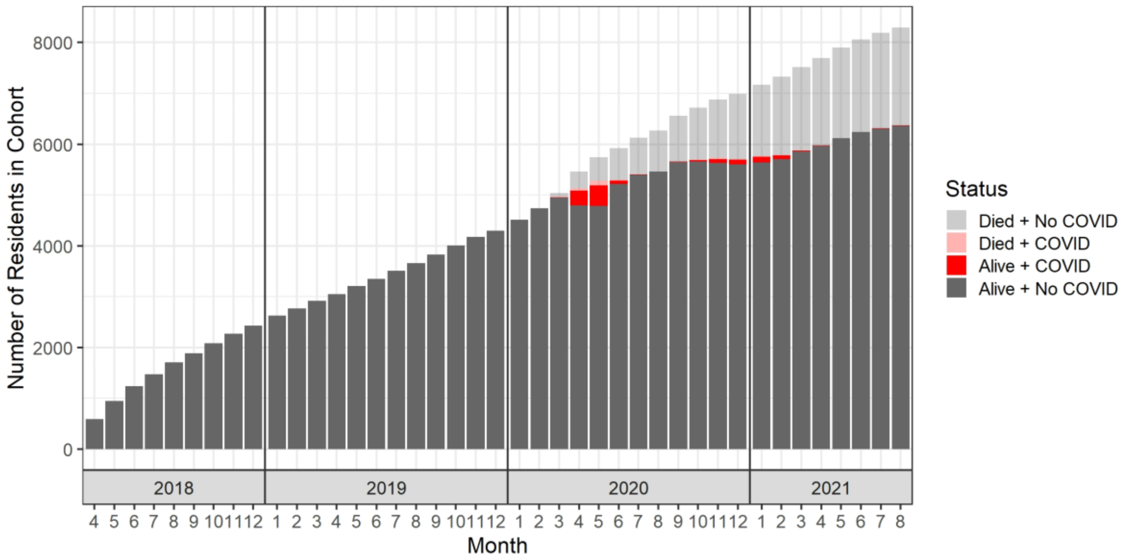


*Figure 2: Number of residents in the cohort over time, also indicating the total number of residents that have died and how many tested positive for COVID-19 each month.*

#### Location and Test Result Correlation

##### *Overall*

We can see the association between where residents have been tested, and the result of the test. We take one test per resident per day and link to their activity on the same day. This results in 14,005 tests. Results indicate that the tests in our dataset are not spread evenly across the locations. The most common location for tests was inpatient with 53% (7,477) of the tests, with A&E the second most common with (23%). The least common was outpatients, with 0.1% (16) of the tests. It also appears that the test result is associated with the location of the test (chi-squared test for independence $p \leq 0.0005$). Tests with a community interaction (both internal and external) on the same date were positive 26% of the time. The rest of the interactions are generally around 5% positive.

The correlations between where residents have been tested, and the result of the test.

*Table 2: Table of proportions of test results in each of the locations where the residents received a COVID-19 test.*

| | Positive | Negative | Not tested | Total |
|---|---|---|---|---|
| Care Home | 6% | 91% | 3% | 2,447 |
| Care Home (Community check up) | 19% | 78% | 3% | 2,156 |
| Community External | 17% | 80% | 3% | 82 |
| A&E | 6% | 92% | 2% | 4,889 |
| Inpatient (Hospital) | 4% | 94% | 3% | 12,842 |
| Outpatient (Hospital) | | 100% | | 28 |

Fisher's exact test for relationship between the locations and test result:

p-value < 0.0005

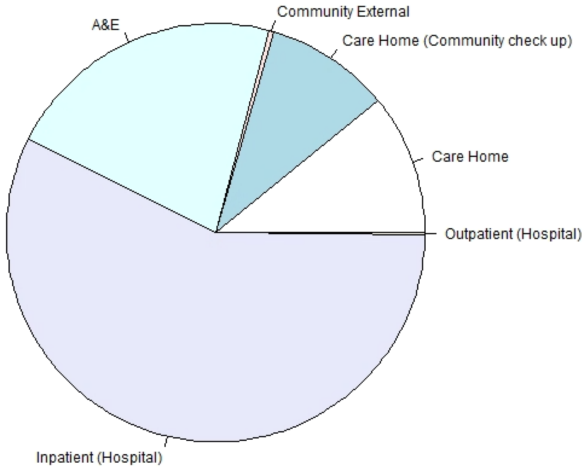


*Figure 3: Locations of COVID-19 tests in the data.*

#### *Wave 1*

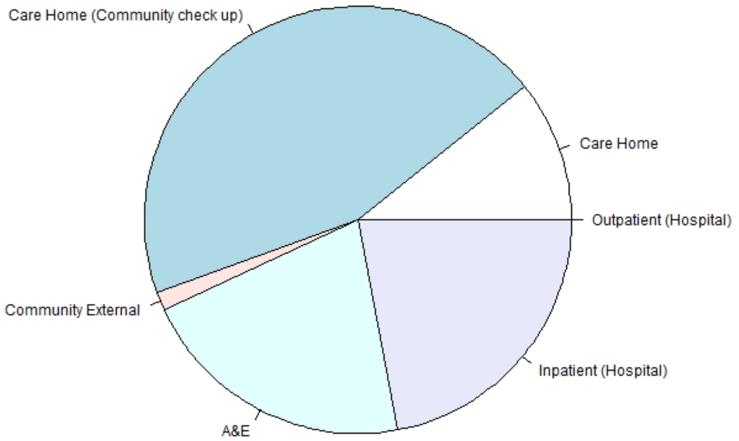


*Figure 4: Locations of tests of care home residents during the first wave.*

*Table 3: COVID-19 tests for care home residents stratified by location of the test and test result for the first wave.*

| | Positive | Negative | No Result | Total |
|---|---|---|---|---|
| Care Home | 56% | 42% | 2% | 155 |
| Care Home (Community check-up) | 53% | 45% | 2% | 641 |
| Community External | 57% | 43% | 0% | 21 |
| A&E | 24% | 72% | 4% | 302 |
| Inpatient (Hospital) | 23% | 74% | 2% | 316 |
| Outpatient (Hospital) | NA | NA | NA | 0 |

***Wave 2***

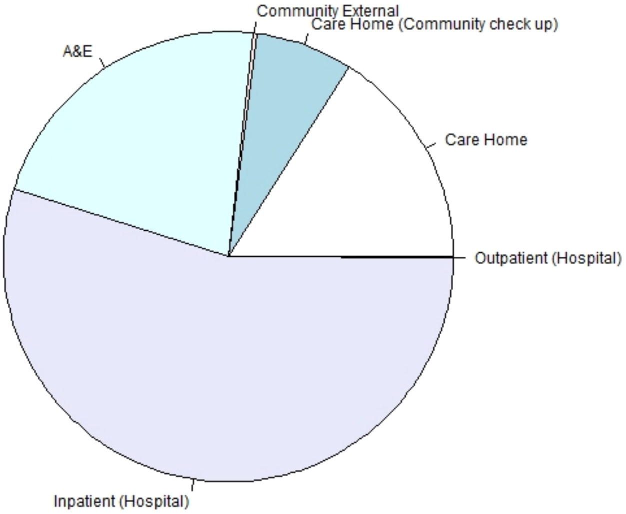


*Figure 5: Locations of COVID-19 tests of care home residents in the second wave.*

*Table 4: COVID-19 tests for care home residents stratified by location of the test and test result for the second wave.*

| | Positive | Negative | No Result | Total |
|---|---|---|---|---|
| Care Home | 3% | 94% | 3% | 1,083 |
| Care Home (Community check-up) | 7% | 99% | 5% | 473 |
| Community External | 0 | 94% | 6% | 17 |
| A&E | 7% | 92% | 2% | 1,482 |
| Inpatient (Hospital) | 4% | 93% | 3% | 3,705 |
| Outpatient (Hosptial) | 0 | 100% | 0 | 7 |

There is a large difference between where the tests are conducted during the first wave and the second wave. The COVID-19 testing in the (pillar 1) data is more often conducted in the care homes during the first wave, whereas the testing is much more common in hospital settings (inpatient and A&E) during the second wave of the pandemic. Overall rates of positive tests in the residents reduce dramatically in the second wave.

#### Cluster Associations Table

Included here are the raw numbers of crossover between characteristics and clusters and p-values of the chi-square tests for independence between the resident characteristics/outcomes and the cluster assignments of the sequences.

*Table 5: Table of p-value of ci-squared test of independence between the cluster assignments and corresponding outcomes and comorbidities.*

| | Mortality | Diabetes | Dementia | Charlson CI |
|---|---|---|---|---|

| | | | | |
|---|---|---|---|---|
| 10 Day Before First Positive | 0.71 | 0.00026 | 0.15 | 0.27 |
| 10 Day After First Positive | 0.12 | 1.0e-08 | 0.00036 | 0.83 |
| 10 Day Before All First Tests | 0.68 | 2.0e-64 | 0.088 | 1.4e-08 |
| 10 Day After All First Tests | 9.3e-08 | 1.1e-24 | 4.7e-17 | 0.0092 |

#### Cluster Associations with Test Result

Associations between test result and the cluster assignments were investigated in order to better understand the testing regime.

| | | Test Result | | Test Result |
|---|---|---|---|---|
| | | Pos | Neg | |
| **10 Days Before First Positive** | Cluster 1 (Inpatient) n = 142 | NA | NA | NA |
| | Cluster 2 (Home) n = 709 | NA | NA | |
| **10 Days After First Positive** | Cluster 1 (Inpatient) n = 195 | NA | NA | NA |
| | Cluster 2 (Home) n = 538 | NA | NA | |
| **10 Days Before All First Tests** | Cluster 1 (Home) n = 3,159 | 81 | 81 | 0.00035 |
| | Cluster 2 (Community) n = 140 | 94 | 94 | |
| | Cluster 3 (Inpatient) n = 172 | 86 | 86 | |
| **10 Days After All First Tests** | Cluster 1 (Inpatient) n = 810 | 92 | 92 | 3.0e-31 |
| | Cluster 2 (Inpatient/Home) n = 578 | 93 | 93 | |
| | Cluster 3 (Home) n = 1,832 | 77 | 77 | |

#### Cluster Associations with Wave of Pandemic

We also tested for associations between wave of the pandemic the test occurred in, and the cluster assignments.

*Table 6: Table of associations between cluster assignments and the wave the index event occured in.*

| | | Wave | | p-value |
|---|---|---|---|---|
| | | 1 | 2 | |
| **10 Days Before First Positive** | Cluster 1 (Inpatient) n = 142 | 22 | 78 | 2.2e-16 |
| | Cluster 2 (Home) n = 709 | 76 | 24 | |
| **10 Days After First Positive** | Cluster 1 (Inpatient) n = 195 | 32 | 68 | 2.2e-16 |
| | Cluster 2 (Home) n = 538 | 80 | 20 | |
| **10 Days Before All First Tests** | Cluster 1 (Home) n = 3,159 | 42 | 58 | 0.016 |
| | Cluster 2 (Community) n = 140 | 29 | 71 | |
| | Cluster 3 (Inpatient) n = 172 | 48 | 52 | |
| **10 Days After All First Tests** | Cluster 1 (Inpatient) n = 810 | 28 | 72 | 8.35e-20 |
| | Cluster 2 (Inpatient/Home) n = 578 | 32 | 68 | |
| | Cluster 3 (Home) n = 1,832 | 49 | 51 | |

Inpatient clusters are more common during Wave 2 of the pandemic, for the sequences both before and after a resident's first positive COVID-19 test. Only a small number of residents' first tests occur within Wave 2, which is likely why the association between wave and cluster assignment before a resident's first test is not significant. However, the cluster assignments after a resident's first test appear to still be statistically significant, in-keeping with the trend of more residents receiving higher levels of care during the second wave.

#### Clustering Waves Separately

##### Comparing Wave 1 and Wave 2 Tests

We look for resident first positive tests and first tests, then filter for the ones during Wave 1 and Wave 2. These are subsets of the overall clustering in the main paper.

**Wave 1**

**Clusters**

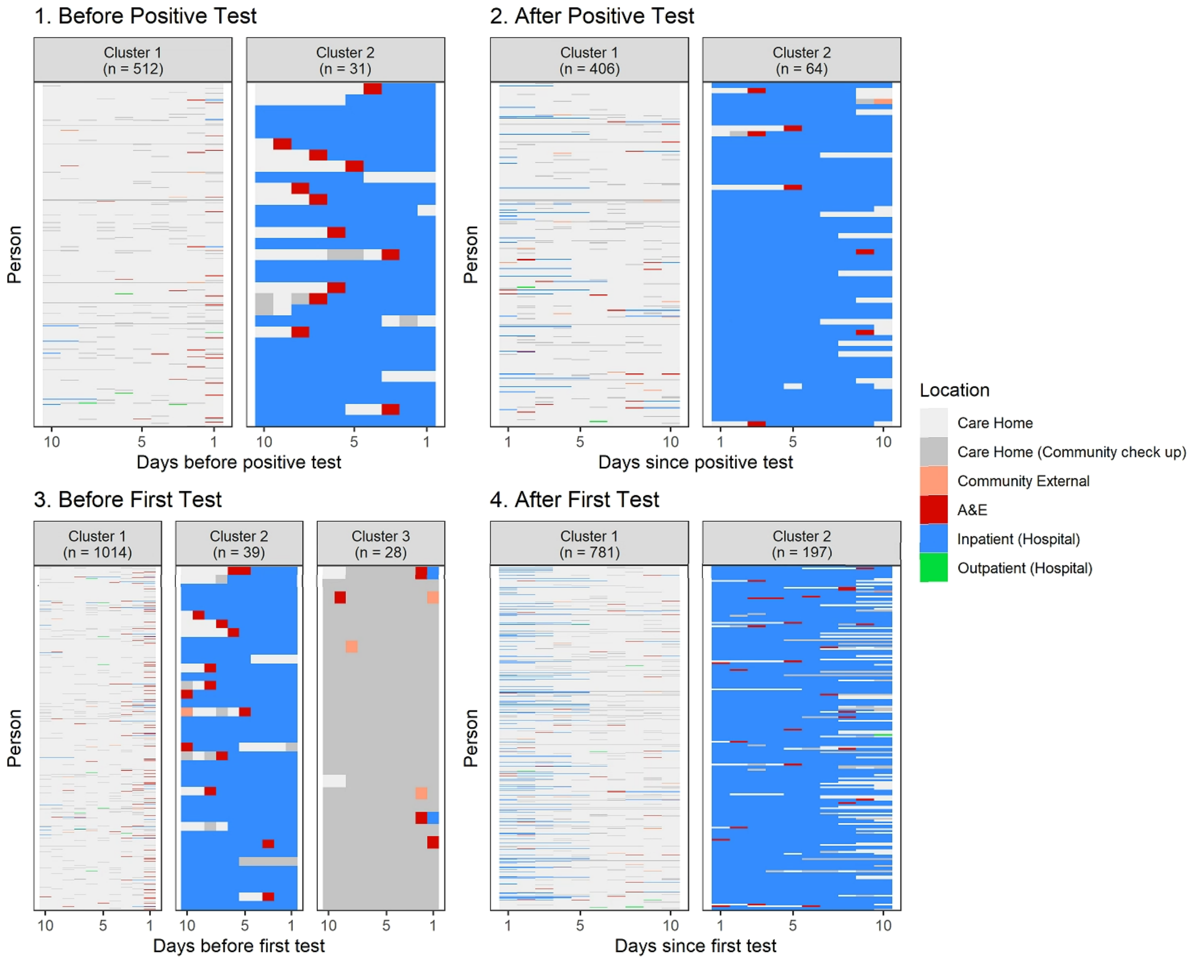


*Figure 6: Cluster assignments describing typical patterns of care before and after residents' first positive tests and first tests during the first wave of the pandemic.*

The clustering here looks similar to the overall one in the main paper. There is a slightly smaller proportion of inpatients in each assignment.

*Table 7: Table of the associations between the cluster assignments of the trajectories before and after a resident's first positive test and a resident's first test, and the characteristics of the residents in the cluster. Filtered for wave 1.*

| | | **28 Day Mortality** | | **Diabetes** | | **Dementia** | | **Charlson CI (Proportion of those with a CCI)** | | | |
|---|---|---|---|---|---|---|---|---|---|---|---|
| | | T (%) | F (%) | T (%) | F (%) | T (%) | F (%) | 0 (%) | 1-2 (%) | 3-4 (%) | ≥5 (%) |
| **10 Day Before First Positive** | Cluster 1 (Inpatient) n = 142 | 23 | 77 | 18 | 82 | 20 | 80 | 08 | 54 | 29 | 09 |
| | Cluster 2 (Home) n = 709 | 23 | 77 | 35 | 65 | 23 | 77 | 00 | 53 | 33 | 13 |
| **10 Day After First Positive** | Cluster 1 (Inpatient) n = 195 | 10 | 90 | 15 | 85 | 18 | 82 | 06 | 54 | 31 | 09 |
| | Cluster 2 (Home) n = 538 | 19 | 81 | 38 | 62 | 33 | 67 | 05 | 55 | 29 | 11 |
| **10 Day Before All First Tests** | Cluster 1 (Home) n = 3,159 | 15 | 85 | 18 | 82 | 20 | 80 | 08 | 52 | 30 | 10 |
| | Cluster 2 (Community) n = 140 | 15 | 85 | 41 | 59 | 21 | 79 | 05 | 37 | 40 | 18 |
| | Cluster 3 (Inpatient) n = 172 | 14 | 86 | 93 | 07 | 46 | 54 | 00 | 08 | 60 | 32 |
| **10 Day After All First Tests** | Cluster 1 (Inpatient) n = 810 | 06 | 94 | 18 | 82 | 20 | 80 | 08 | 50 | 31 | 12 |
| | Cluster 2 (Inpatient/Home) n = 578 | 07 | 93 | 32 | 68 | 25 | 75 | 06 | 47 | 36 | 11 |
| | Cluster 3 (Home) n = 1,832 | 23 | 77 | 18 | 82 | 20 | 80 | 08 | 54 | 29 | 09 |

*Table 8: Table of the p-values of the associations between the cluster assignments of the trajectories before and after a resident's first positive test and a resident's first test, and the characteristics of the residents in the cluster. Filtered for wave 1.*

| | Mortality | Diabetes | Dementia | Charlson CI |
|---|---|---|---|---|
| 10 Day Before First Positive | 0.95 | 0.017 | 0.76 | 0.93 |
| 10 Day After First Positive | 0.058 | 4.1e-05 | 0.013 | 0.96 |
| 10 Day Before All First Tests | 1 | 1.1e-22 | 0.0029 | 5.7e-0.6 |
| 10 Day After All First Tests | 0.72 | 8.9e-06 | 0.07 | 0.63 |

The cluster assignments for the testing in the first wave and characteristics of the residents generally have fewer significant associations than the full clustering. This is likely due to the smaller sample size. Diabetes is generally significantly associated with the higher care consumption clusters as seen previously, however the before the first positive test this is not the case. The percentage difference looks to fit the trend, however, is not large enough to be significant.

Half of the residents in the care home after their test, tested positive, while only 24% of the residents who were an inpatient after their test were positive.

Dementia was not found to be significant for any of the cluster assignments, indicating residents with dementia were significantly treated in one particular.

Charlson comorbidity index was found to be significant for the care before first COVID-19 test, with residents with a higher index being more often in higher levels of care. This is, likely due to the presence of the inpatient cluster. The residents in hospital were also frailer than those who stayed in the home.

**Wave 2**

**Clusters**

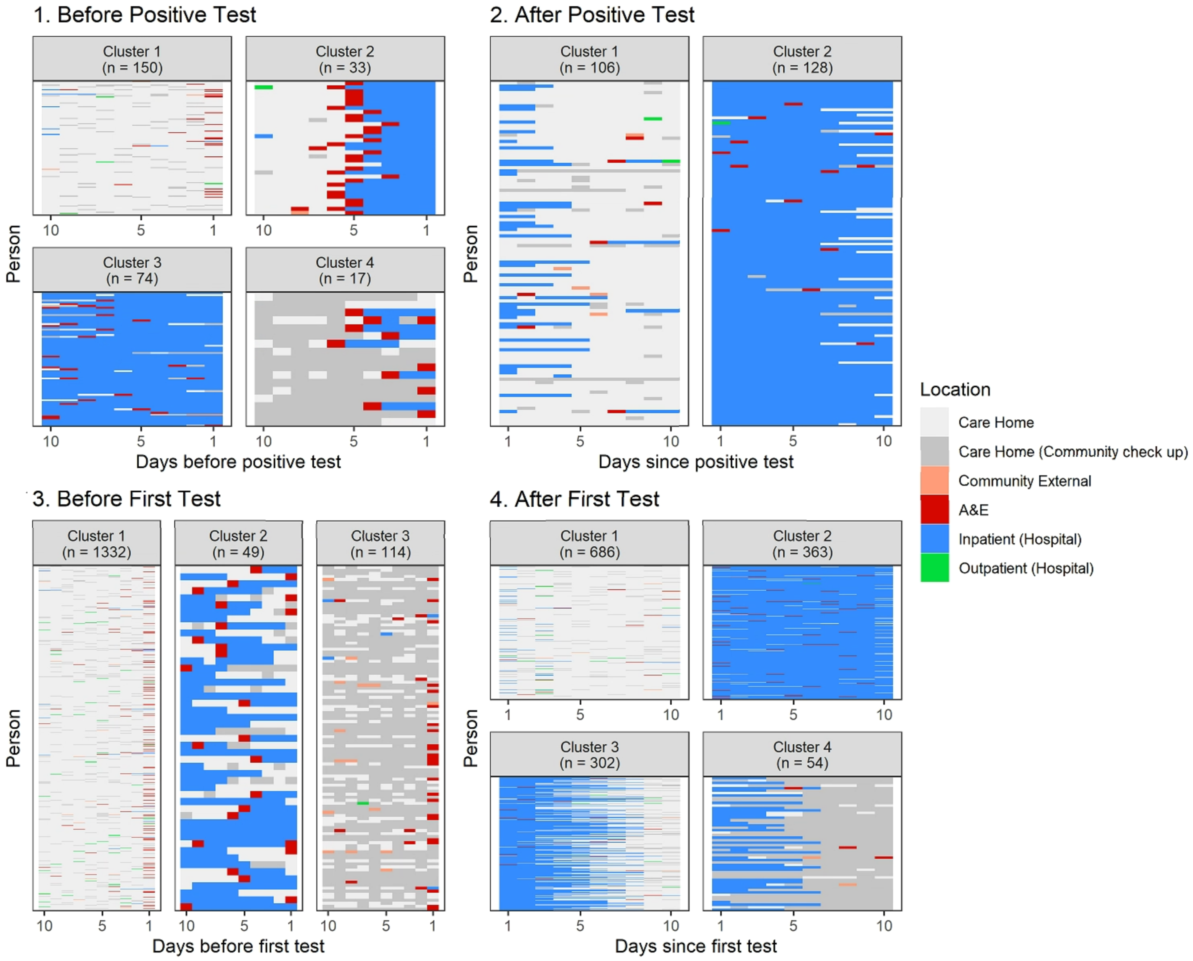


*Figure 7: Cluster assignments describing typical patterns of care before and after residents' first positive tests and first tests during the second wave of the pandemic.*

| | | 28 Day Mortality | | Diabetes | | Dementia | | Charlson CI (Proportion of those with a CCI) | | | |
|---|---|---|---|---|---|---|---|---|---|---|---|
| | | T (%) | F (%) | T (%) | F (%) | T (%) | F (%) | 0 (%) | 1-2 (%) | 3-4 (%) | ≥5 (%) |
| **10 Day Before First Positive** | Cluster 1 (Inpatient) n = 142 | 26 | 74 | 26 | 74 | 24 | 76 | 52 | 05 | 43 | 00 |
| | Cluster 2 (Home) n = 709 | 18 | 82 | 27 | 73 | 33 | 67 | 52 | 14 | 27 | 07 |
| **10 Day After First Positive** | Cluster 1 (Inpatient) n = 195 | 24 | 76 | 39 | 61 | 26 | 74 | 49 | 13 | 31 | 07 |
| | Cluster 2 (Home) n = 538 | 12 | 88 | 59 | 41 | 24 | 76 | 32 | 26 | 42 | 00 |
| **10 Day Before All First Tests** | Cluster 1 (Home) n = 3,159 | 09 | 91 | 25 | 75 | 18 | 82 | 12 | 44 | 38 | 07 |
| | Cluster 2 (Community) n = 140 | 12 | 88 | 35 | 65 | 30 | 70 | 07 | 46 | 36 | 11 |
| | Cluster 3 (Inpatient) n = 172 | 11 | 89 | 20 | 80 | 22 | 79 | 08 | 53 | 32 | 07 |
| **10 Day After All First Tests** | Cluster 1 (Inpatient) n = 810 | 10 | 90 | 35 | 65 | 08 | 92 | 06 | 57 | 30 | 06 |
| | Cluster 2 (Inpatient/Home) n = 578 | 11 | 89 | 56 | 44 | 22 | 78 | 09 | 35 | 41 | 15 |
| | Cluster 3 (Home) n = 1,832 | 01 | 98 | 15 | 85 | 14 | 86 | 09 | 50 | 35 | 06 |

The '*After positive test*' sequence definition in the second wave is the only set where the care home cluster is not the most common cluster. This reflects the fact the pillar 1 testing in the second wave is more routine testing on arrival to hospital. The inpatient clusters include a larger proportion of the residents in the '*After*' sequences than in the overall testing clusters, evidencing this further.

*Table 9: Table demonstrating the associations between the cluster assignments of the trajectories before and after a resident's first positive test and a resident's first test, and the characteristics of the residents in the cluster. Filtered for wave 2.*

| | Mortality | Diabetes | Dementia | Charlson CI |
|---|---|---|---|---|
| 10 Day Before First Positive | 0.56 | 0.44 | 0.72 | 0.44 |
| 10 Day After First Positive | 0.73 | 0.11 | 0.05 | 0.67 |
| 10 Day Before All First Tests | 0.99 | 1. 0e-17 | 0.079 | 0.016 |
| 10 Day After All First Tests | 4.9e-04 | 1.7e-13 | 4.0e-08 | 0.21 |

There appears to be no statistically significant relationship between diabetes and the clusters for the sequences before and after a resident's first positive test. This is also likely due to the sample size, as we still see the residents with diabetes in the clusters relating to higher levels of care.

After a resident's first test, residents with dementia make up 31% of the community cluster and make up a similar proportion of residents in the two inpatient clusters and are more prevalent than they are in the home cluster.

From the tests in the second wave, residents who have been an inpatient after their test have the highest rate of deaths. This is intuitive, since residents receiving the highest levels of care are likely to be the highest mortality risk.

##### Limitations of State Sequence Analysis

State sequence analysis in this application quantises care into discrete states, with one per day-collapsing down any days where more than one event occurs. We use a resolution of 10 days that attempts to balance complexity/length of the sequences and how well it represents the events happening during the sequence. Smaller time units would allow a more precise description of events but can result in sequences and clusters that are to interpret over longer time periods. Additional contextual information is also not included in the analysis, so specific circumstances/reasons for each healthcare event are not included.

The sequences are treated as a whole, where patterns of states are identified. A transition matrix is used to define the substitution costs through the transition rates between the states, however order forwards and backwards in the sequences are treated equally. The sequence analysis does not have a temporal component other than the order of the sequence, and therefore transitions backwards are treated equally to transitions forwards.

#### References


1. Department of Health and Social Care. Coronavirus (COVID-19) - Scaling up our testing programmes. Published online April 4, 2020. https://assets.publishing.service.gov.uk/government/uploads/system/uploads/attachment_data/file/878121/coronavirus-covid-19-testing-strategy.pdf

2. Abbott A, Forrest J. Optimal Matching Methods for Historical Sequences. *The Journal of Interdisciplinary History*. 1986;16(3):471-494. doi:10.2307/204500

3. Needleman SB, Wunsch CD. A general method applicable to the search for similarities in the amino acid sequence of two proteins. *J Mol Biol*. 1970;48(3):443-453. doi:10.1016/0022-2836(70)90057-4
